# Internal consistency and concurrent validity of the Peruvian Infant Development Scale tool in high-altitude rural Andean Peru

**DOI:** 10.1101/2024.06.09.24308664

**Authors:** Néstor Nuño, Daniel Mäusezahl, Dana McCoy, Stella María Hartinger

## Abstract

**Background:** Measuring the developmental status of children living in low- and middle-income countries is a major challenge. The Peruvian Infant Development Scale (ESDI) is a screening instrument used for children under 36 months of age. The ESDI has been validated in Peru, but its psychometric properties have not been yet compared with international instruments.

**Methods:** In rural Andean Peru, we compared ESDI scores with those of the Bayley Scales of Infant Toddler Development (BSID) to investigate the internal consistency and concurrent validity of the ESDI. Internal consistency was assessed using standardised Cronbach’s alpha coefficients (α) and item-test correlations. We examined the concurrent validity of cognitive, motor, and communication domains using Pearson correlations (*r*) and bootstrapping methods in children aged 19-30 months and 31-36 months. We also examined the relationship between the developmental domains, maternal education, and household wealth.

**Results:** We compared information from 167 children. Internal consistency and reliability were acceptable for both the ESDI (α=0.75; item-test: 0.68-0.85) and the BSID (α=0.91; item-test: 0.82-0.90). In children aged 19-30 months, we found weak correlations for the cognitive (*r*=0.04; *p*>0.05) and motor (*r*=-0.04-0.11; *p*>0.05) domains, and moderate-to-strong correlations for the communication domain (*r*=0.42-0.57; *p*<0.05). At 31-36 months of age, we found moderate correlations for the cognitive (*r*=0.46; *p*<0.05) and communication (*r*=0.37-0.44; *p*<0.05) domains, and weak-to-moderate correlations for the motor domain (*r*=0.22-0.31; *p*<0.05). The ESDI and BSID domains were equally correlated with maternal education and household wealth.

**Conclusions:** The ESDI is a useful instrument for assessing the developmental status of children, especially those over 30 months of age. Due to its free cost, short duration, and simple administration, the ESDI is suitable for resource-poor settings in Peru. The ESDI can be strategically scaled through the Peruvian ECD programme *Cuna Mas*, so that this intervention can be adequately monitored in a cost-effective manner.

## 1. Introduction

In the early stages of childhood, humans acquire the abilities to move, coordinate, think, feel, and interact with others and their environment (Schneider & Ramires, 2008; Wachs, 2000). The early childhood period begins during pregnancy and continues through the first three years of life (United Nations Children’s Fund, 2012). The experiences and skills acquired in childhood are critical to the development of learning abilities, behaviours, and physical and mental health throughout life (S Grantham-McGregor & Smith, 2016; Hoffman, 2008).

A number of factors contribute to good child development, including health, nutrition, safety, responsive parental care, and early stimulation (Black et al., 2017; Britto, Engle, & Super, 2013). In low- and middle-income countries (LMICs), children often receive less stimulation during their early childhood years than those in high-income countries (Bornstein & Putnick, 2012; S Grantham-McGregor et al., 2007). In 2010, approximately 249 million under-five children were at risk of not reaching their full cognitive potential because they were undernourished or extremely poor (Lu, Black, & Richter, 2016). Studies have shown that inequalities in early childhood have a negative effect on the long-term health, development, educational attainment, and well-being of future generations (Shonkoff & Richter, 2013; Walker et al., 2011).

In Peru, children from different income quartiles and geographical regions show large differences in development (Schady et al., 2015). In a recent study of rural Peruvian Amazonian children aged 8-38 months, a quarter were found to have developmental delays (Westgard & Alnasser, 2017). Another study with 1,176 under-five children from poor areas of Peru found that 47% had difficulty reaching at least one age-specific motor milestone and 53% had difficulty reaching at least one age-specific verbal milestone. Children from households with poor housing infrastructure, overcrowding, or lack of access to safe drinking and adequate sanitation and whose mothers had a low level of education showed the worst developmental outcomes (Díaz, Gallestey, Vargas-Machuca, & Velarde, 2017).

Various studies have shown that children from disadvantaged backgrounds who received early child development (ECD) interventions are more likely to achieve academic success, be socially and emotionally stable, and be economically productive in adulthood (Gertler et al., 2014; S. Grantham-McGregor, 2015; Woodhead, Ames, & Streuli, 2009). A number of systematic reviews and meta-analysis also confirmed the effectiveness of ECD interventions for improving the developmental status of children in LMICs when implemented by caregivers (Jeong, Franchett, Ramos de Oliveira, Rehmani, & Yousafzai, 2021) and healthcare providers (Hirve et al., 2023). For these reasons, ECD interventions are considered one of the most cost-effective strategies for reducing inequalities in LMIC (Engle et al., 2011; Irwin, Siddiqi, & Hertzman, 2007).

In recent decades, various measures have been used to assess the developmental status of Peruvian children. However, the application of these instruments has proven difficult in rural areas with limited human and logistical resources, as many of them require professionals to administer and evaluate them (Gutierrez, Lazarte, & Alarcon, 2016). The Peruvian Infant Development Scale (“*Escala de Desarrollo Infantil*” (ESDI)) has overcome this hurdle as it is a screening tool that can be easily used in a variety of cultural contexts by non-professionals (Programa Nacional Cuna Mas, 2016).

The content and reproducibility of the ESDI have been evaluated in the past (Programa Nacional Cuna Mas, 2016), but the congruence of the ESDI to correctly measure child development outcomes has not yet been compared with other similar instruments. The lack of reliable and valid instruments to measure child development outcomes can hinder the success of ECD interventions (Munoz-Chereau, Ang, Dockrell, Outhwaite, & Heffernan, 2021). This study aims to assess the internal consistency and concurrent validity of the ESDI in the high-altitude rural Andes of Peru. Our study compares ESDI scores with those of the Bayley Scales of Infant Toddler Development (BSID) (Bayley, 2005), the gold standard for measuring children’s developmental status.

## 2. Methods

### 2.1. Study site and participants

The study was conducted in the provinces of San Marcos and Cajabamba, two high-altitude rural resource-limited locations in the northern highlands of Peru (Cajamarca region). The participants in this study were among the 317 children from 82 rural communities who participated in a cluster-randomised controlled trial to evaluate the Peruvian national ECD programme (“*Programa Nacional Cuna Más*” (PNCM)). Children belonged to families with low income and education level and lived in households with adobe walls and limited access to safe drinking water and adequate sanitation (Hartinger, Nuño, Verastegui, Ortiz, & Mäusezahl, 2020).

### 2.2. Study design and data collection

ECD information was collected from all participants at the end of the trial. Four fieldworkers, trained by PNCM experts for one week, applied the ESDI tool between April and July 2017. Subsequently, a team of specialist psychologists from the University Cayetano Heredia (“*Universidad Peruana Cayetano Heredia*” (UPCH)) administered the BSID to the same children between June and September 2017. Both instruments were used and evaluated according to the respective manuals.

### 2.3. Instruments

The ESDI is suitable for children under 36 months of age and assesses four developmental domains: cognitive, communication, motor, and socio-emotional skills. The items are rated on a two-point scale (0-100 points per domain). In addition, the ESDI provides and overall developmental score, which is calculated from the average of the scores achieved in the domains. Most items are scored through direct structured observation, although caregivers may report some of them. The ESDI is free, takes about 90 minutes to complete, those administering it only need to have a secondary school diploma, and the required training takes about one week.

In 2016, the psychometric properties of the ESDI were examined with 127 children from 11 Peruvian regions. The test-retest comparisons confirmed the reproducibility of the instrument. The Pearson coefficients (*r*) for cognitive skills were *r*=0.87, *r*=0.82 for communication skills, *r*=0.88 for motor skills, *r*=0.81 for socio-emotional skills, and *r*=0.90 for overall development. In addition, the ESDI was reviewed by a panel of experts to validate its content (e.g., wording and age-related adaptations) (Programa Nacional Cuna Mas, 2016).

The BSID assesses the development of children under 42 months of age in five developmental domains: cognitive, receptive language, expressive language, fine motor, and gross motor skills. The items consist of a series of play-tasks that are scored through direct observation by the examiner on a two-point scale. The raw BSID scores can be converted to a 1-19 scale (mean=10, SD=3), and 40-160 composite scale (mean=100, SD=15), which is compared to a standardised sample from the United States. In the BSID composite scale, fine and gross motor skills are combined into a single category. The same applies to receptive and expressive language. The BSID costs about 1,025US$ per kit, takes about 120 minutes to complete, and those administering it must have a degree in psychology and about one month of specialised formal training. In this study, we used the Spanish adaptation of the BSID, which was recently validated for Peru (Murray-Kolb et al., 2014; Pendergast et al., 2018).

### 2.4. Data analysis

We conducted a psychometric analysis using the classical test theory (Alagumalai & Curtis, 2005). To determine the effect of age, the analysis was stratified into two age groups: 19-30 and 31-36 months. These age groups have been previously used in similar studies (Rubio-Codina, Araujo, Attanasio, Muñoz, & Grantham-McGregor, 2016). Children with ESDI information who did not participate in or complete the BSID assessments, as well as children who completed the ESDI in one age group and changed age groups at the time of the BSID, were excluded from the analysis. We only used raw BSID scores, as there were no norms adapted to the local context.

We used standardised Cronbach’s alpha coefficients (α) and item-test correlations to assess the internal consistency of the ESDI and BSID instruments. The item-test correlation describes the relationship between the individual items and the overall scale. In general, a higher item-test correlation score indicates better internal consistency results, as it indicates how positively the items contribute to the instrument (Salkind, 2010). We defined good internal consistency as Cronbach’s alpha coefficients ≥0.70 and item-test correlations ≥0.40.

We assessed the concurrent validity of the ESDI tool using Pearson correlations and bootstrapping methods of 1,000 replications adjusted for correlation within cluster. We compared ESDI and BSID domains that measure similar developmental outcomes. The ESDI domain communication was compared to the BSID domains expressive language and receptive language. We also compared the ESDI domain motor to the BSID domains fine motor and gross motor. The socio-emotional domain of the ESDI was excluded, as there was no matching BSID domain. We classified Pearson correlations as weak (*r*≤0.29), moderate (*r*=0.30-0.49), and strong (*r*≥0.50) (Piedmont, 2014). We defined good concurrent validity as positive and moderate correlations. To further investigate concurrent validity, we used generalised estimated equation (GEE) models with robust standard errors and an independent correlation structure to assess the association between the ESDI and BSID domains, female education (in years), and household wealth. This structure provides robust results regardless of the number of participants in the different clusters. Coefficients were controlled for child’s age, child’s sex, and within-cluster correlation. Adjustment variables were selected based on similar studies (Rubio-Codina et al., 2016; Yue et al., 2019). We classified the wealth of households using the Young Lives Wealth Index. This index classifies household wealth based on three indicators: housing quality (e.g., wall and roof materials), access to services (e.g., electricity and fuel for cooking), and ownership of consumer durables (e.g., radio, and refrigerator). All three indicators are scored equally and the overall wealth index (between 0 and 1) is calculated by taking the average of the three indices. A higher value indicates a higher socio-economic status (Briones, 2017). The statistical analysis was performed using Stata 15.1 (StataCorp, College Station, Texas) and R 3.4 (R Foundation for Statistical Computing).

### 2.5. Ethics

The ethics committees of the regional health authority of Cajamarca and the UPCH approved the study in which this trial is embedded (Ref. 268-12-15). Participants’ families signed an informed consent form prior to participation. All information received was treated confidentially. The study was registered with ISRCTN (study register: ISRCTN26548981).

## 3. Results

ESDI information was collected from 204 children. Eleven children with ESDI assessments who did not participate in the BSID assessment and 26 children, who completed ESDI and BSID assessments at different times so that they moved to a different age group, were excluded from the analysis. The final analysis included information from 167 children – 80 aged 19-30 months and 87 aged 31-36 months. Figure 1 shows a detailed flowchart of participants.

**Figure 1.**
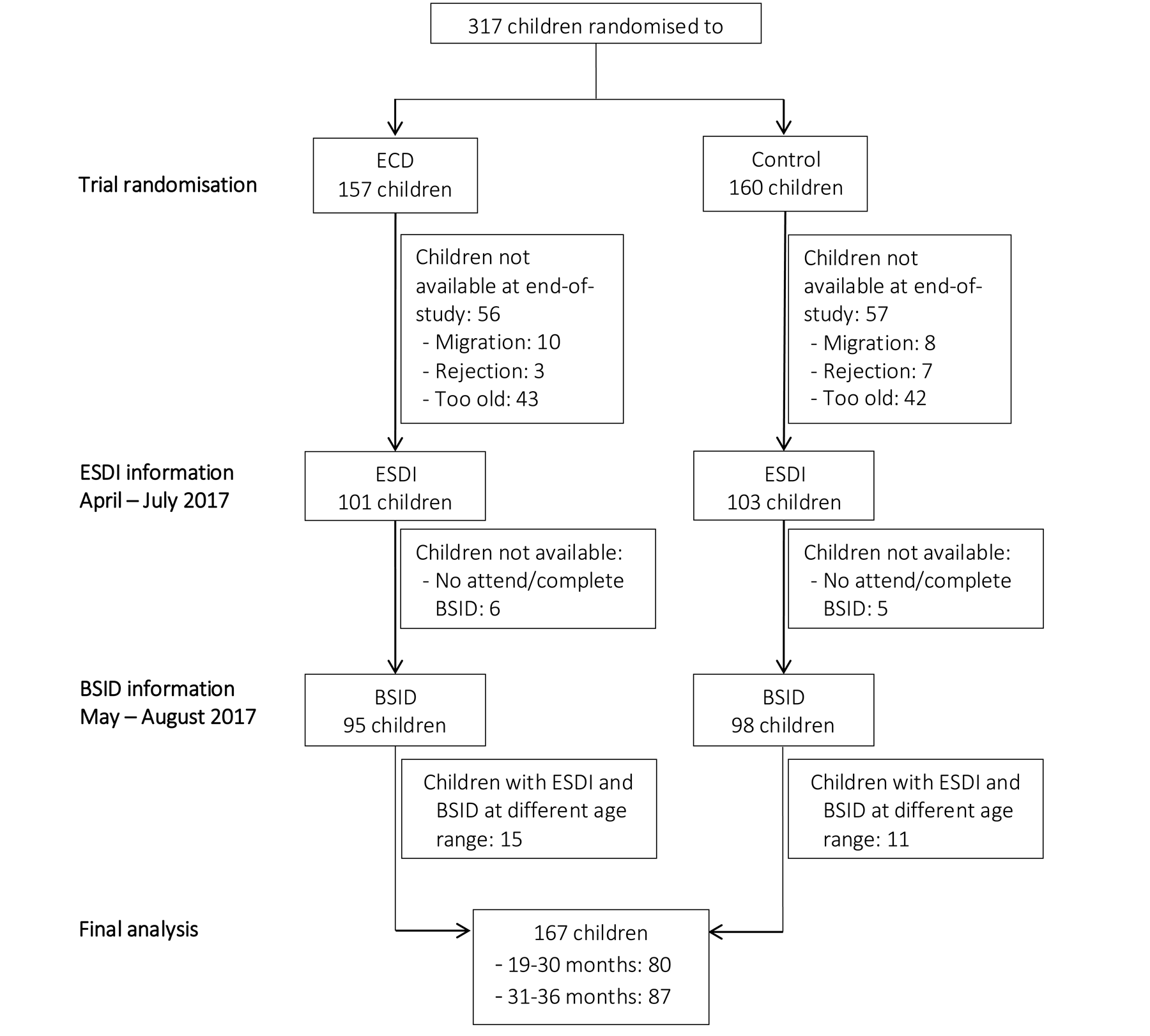
Flowchart of study participants

### 3.1. Participant characteristics

A total of 53% (N=86) of the children were female and the average birthweight was 3.1 (SD=0.5) kilogrammes. The average age of male and female caregivers was 29.1 (SD=7.5) and 33.9 (SD=8.2) years, respectively. The time difference between ESDI and BSID assessments was 54.4 (SD=33.8) days.

**Table 1.**
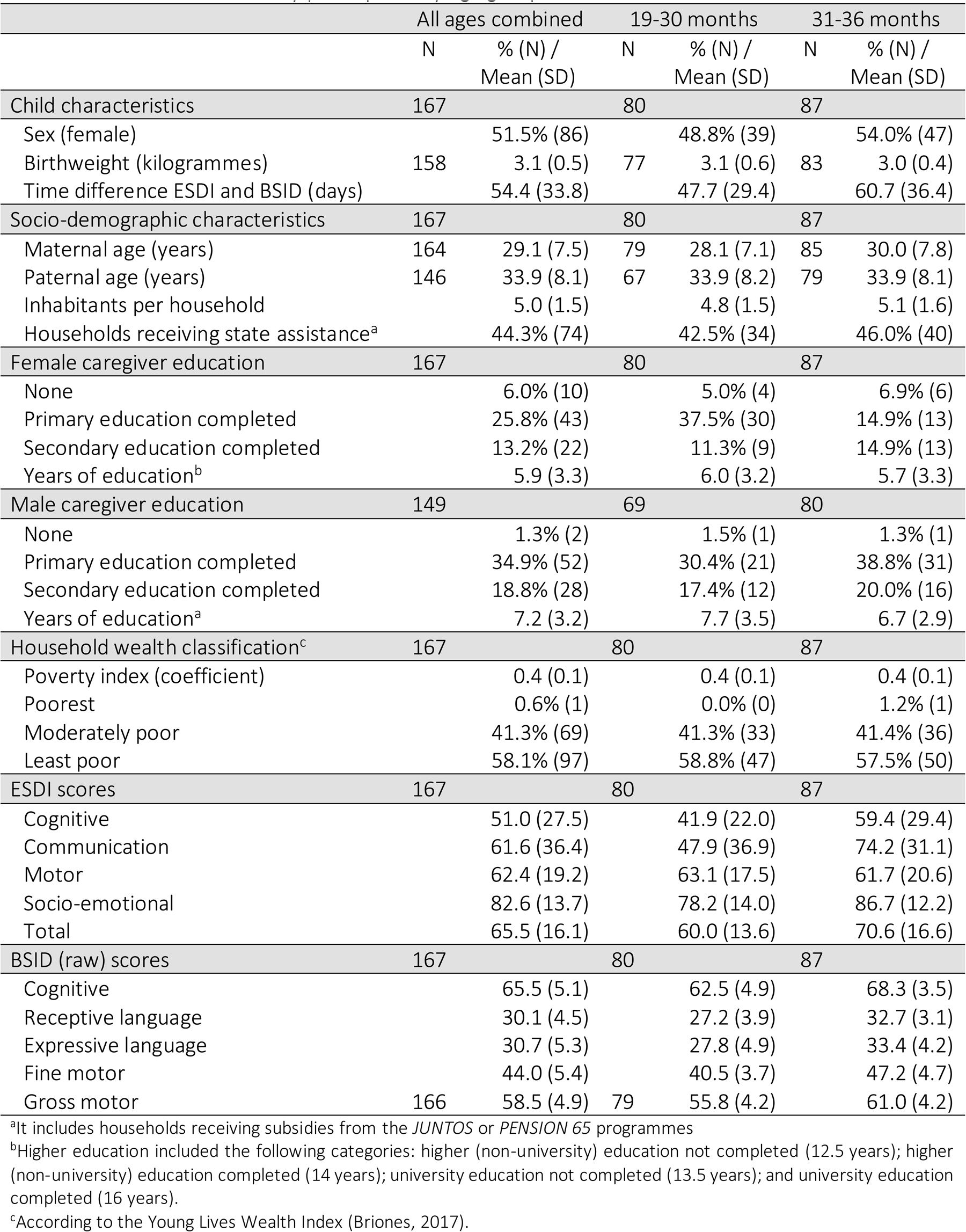
Characteristics of study participants by age group.

|  | All ages combined |  | 19-30 months |  | 31-36 months |  |
| --- | --- | --- | --- | --- | --- | --- |
|  | N | % (N) /<br>Mean (SD) | N | % (N) /<br>Mean (SD) | N | % (N) /<br>Mean (SD) |
| **Child characteristics** | **167** |  | **80** |  | **87** |  |
| Sex (female) |  | 51.5% (86) |  | 48.8% (39) |  | 54.0% (47) |
| Birthweight (kilogrammes) | 158 | 3.1 (0.5) | 77 | 3.1 (0.6) | 83 | 3.0 (0.4) |
| Time difference ESDI and BSID (days) |  | 54.4 (33.8) |  | 47.7 (29.4) |  | 60.7 (36.4) |
| **Socio-demographic characteristics** | **167** |  | **80** |  | **87** |  |
| Maternal age (years) | 164 | 29.1 (7.5) | 79 | 28.1 (7.1) | 85 | 30.0 (7.8) |
| Paternal age (years) | 146 | 33.9 (8.1) | 67 | 33.9 (8.2) | 79 | 33.9 (8.1) |
| Inhabitants per household |  | 5.0 (1.5) |  | 4.8 (1.5) |  | 5.1 (1.6) |
| Households receiving state assistance<sup>a</sup> |  | 44.3% (74) |  | 42.5% (34) |  | 46.0% (40) |
| **Female caregiver education** | **167** |  | **80** |  | **87** |  |
| None |  | 6.0% (10) |  | 5.0% (4) |  | 6.9% (6) |
| Primary education completed |  | 25.8% (43) |  | 37.5% (30) |  | 14.9% (13) |
| Secondary education completed |  | 13.2% (22) |  | 11.3% (9) |  | 14.9% (13) |
| Years of education<sup>b</sup> |  | 5.9 (3.3) |  | 6.0 (3.2) |  | 5.7 (3.3) |
| **Male caregiver education** | **149** |  | **69** |  | **80** |  |
| None |  | 1.3% (2) |  | 1.5% (1) |  | 1.3% (1) |
| Primary education completed |  | 34.9% (52) |  | 30.4% (21) |  | 38.8% (31) |
| Secondary education completed |  | 18.8% (28) |  | 17.4% (12) |  | 20.0% (16) |
| Years of education<sup>a</sup> |  | 7.2 (3.2) |  | 7.7 (3.5) |  | 6.7 (2.9) |
| **Household wealth classification<sup>c</sup>** | **167** |  | **80** |  | **87** |  |
| Poverty index (coefficient) |  | 0.4 (0.1) |  | 0.4 (0.1) |  | 0.4 (0.1) |
| Poorest |  | 0.6% (1) |  | 0.0% (0) |  | 1.2% (1) |
| Moderately poor |  | 41.3% (69) |  | 41.3% (33) |  | 41.4% (36) |
| Least poor |  | 58.1% (97) |  | 58.8% (47) |  | 57.5% (50) |
| **ESDI scores** | **167** |  | **80** |  | **87** |  |
| Cognitive |  | 51.0 (27.5) |  | 41.9 (22.0) |  | 59.4 (29.4) |
| Communication |  | 61.6 (36.4) |  | 47.9 (36.9) |  | 74.2 (31.1) |
| Motor |  | 62.4 (19.2) |  | 63.1 (17.5) |  | 61.7 (20.6) |
| Socio-emotional |  | 82.6 (13.7) |  | 78.2 (14.0) |  | 86.7 (12.2) |
| Total |  | 65.5 (16.1) |  | 60.0 (13.6) |  | 70.6 (16.6) |
| **BSID (raw) scores** | **167** |  | **80** |  | **87** |  |
| Cognitive |  | 65.5 (5.1) |  | 62.5 (4.9) |  | 68.3 (3.5) |
| Receptive language |  | 30.1 (4.5) |  | 27.2 (3.9) |  | 32.7 (3.1) |
| Expressive language |  | 30.7 (5.3) |  | 27.8 (4.9) |  | 33.4 (4.2) |
| Fine motor |  | 44.0 (5.4) |  | 40.5 (3.7) |  | 47.2 (4.7) |
| Gross motor | 166 | 58.5 (4.9) | 79 | 55.8 (4.2) |  | 61.0 (4.2) |

<sup>a</sup>It includes households receiving subsidies from the *JUNTOS* or *PENSION 65* programmes
<sup>b</sup>Higher education included the following categories: higher (non-university) education not completed (12.5 years); higher (non-university) education completed (14 years); university education not completed (13.5 years); and university education completed (16 years).
<sup>c</sup>According to the Young Lives Wealth Index (Briones, 2017).

The average number of inhabitants per household was 5.0 (SD=1.5). Forty-four percent of households (N=74) received subsidies from the Peruvian national social programmes, *JUNTOS* or *Pension65*. Twenty-six percent (N=43) of female caregivers and 35% (N=52) of male caregivers had completed primary education. This gender difference was also observed among those who had completed secondary education (13% of female *versus* 19% of male caregivers). Some 6% of female caregivers had no education, compared to 1% of male caregivers. The average years of education for female and male caregivers was 5.9 (SD=3.3) and 7.2 (SD=3.2) respectively. According to the Young Lives Wealth Index, the average coefficient was 0.4 (SD=0.1), 41% (N=69) of households were classified as least poor, and 58% as moderately poor. Only one household in the age group of 31-36 months belonged to the lowest socio-economic level.

The lowest and highest average ESDI scores were observed in the cognitive and socio-emotional areas respectively. On the other hand, the lowest BSID scores were observed in both the receptive and expressive language domains, while the cognitive domain had the highest scores. Age groups were balanced in terms of the child’s sex, birthweight, socio-demographic characteristics, fatheŕs education, and household wealth classification. More mothers in the group of 19-30 months had completed elementary school. The children aged 31-36 months showed more time differences between the ESID and BSID assessments and scored higher in all ESDI and BSID domains (Table 1).

### 3.2. Internal consistency and reliability

Table 2 describes the internal consistency and reliability of the ESDI and BSID. The Cronbach’s alpha coefficient for the ESDI (all ages combined) was 0.75, and did not improve when domains were removed. The Cronbach’s alpha coefficients decreased at 19-30 months (α=0.65) and increased at 31-36 months (α=0.80) compared to all ages combined. The item-test correlation values were between 0.62-0.80 (19-30 months), 0.71-0.86 (31-36 months) and 0.68-0.85 (all ages combined).

**Table 2.** Internal consistency of the ESDI and BSID by age group.

| | Item-test correlation | | | Cronbach's alpha ($\alpha$)<sup>a</sup> | | |
| --- | --- | --- | --- | --- | --- | --- |
|  | All ages combined | 19-30 months | 31-36 months | All ages combined | 19-30 months | 31-36 months |
| **ESDI** |  |  |  | **0.75** | **0.65** | **0.80** |
| Cognitive | 0.85 | 0.80 | 0.86 | 0.59 | 0.46 | 0.69 |
| Communication | 0.79 | 0.74 | 0.81 | 0.66 | 0.53 | 0.73 |
| Motor | 0.68 | 0.63 | 0.78 | 0.75 | 0.65 | 0.76 |
| Socio-emotional | 0.69 | 0.62 | 0.71 | 0.74 | 0.65 | 0.80 |
| **BSID** |  |  |  | **0.91** | **0.86** | **0.84** |
| Cognitive | 0.89 | 0.83 | 0.83 | 0.89 | 0.82 | 0.79 |
| Receptive language | 0.90 | 0.85 | 0.80 | 0.90 | 0.81 | 0.81 |
| Expressive language | 0.84 | 0.78 | 0.75 | 0.90 | 0.84 | 0.73 |
| Fine motor | 0.88 | 0.85 | 0.77 | 0.89 | 0.87 | 0.82 |
| Gross motor | 0.82 | 0.69 | 0.76 | 0.91 | 0.69 | 0.82 |

<sup>a</sup>Values in each domain represents Cronbach's alpha coefficient if the domain were removed.

The Cronbach’s alpha coefficient for the BSID (all ages combined) was 0.91 and did not improve when one domain was removed. The Cronbach’s alpha coefficients decreased at 19-30 months (α=0.86) and 31-36 months (α=0.84) compared to all ages combined.

The item-test correlation values were between 0.69-0.85 (19-30 months), 0.75-0.83 (31-36 months) and 0.82-0.90 (all ages combined).

### 3.3. Concurrent validity

Figure 2 shows the correlation matrix between ESDI and BSID (raw) scores by age group. At 19-30 months, we observed weak correlations between the cognitive domains of the ESDI and BSID (*r*=0.04), the ESDI domain motor and the BSID domain fine motor (*r*=-0.04), and the ESDI domain motor and the BSID domain gross motor (*r*=0.11). Communication was the only developmental domain with good agreement in this age group. The correlation between the ESDI domain communication and BSID domain receptive language was moderate (*r*=0.42; *p*<0.001), and the correlation between the ESDI domain communication and the BSID domain expressive language was strong (*r*=0.57; *p*<0.001).

The agreement between the ESDI and BSID improved in the 31 -36 months age group. We observed moderate correlations between the cognitive domains of the ESDI and BSID (*r*=0.46; *p*<0.001), the ESDI domain communication and the BSID domain receptive communication (*r*=0.37; *p*<0.01), the ESDI domain communication and the BSID domain expressive language (*r*=0.44; *p*<0.001), and the ESDI domain motor and the BSID domain gross motor (*r*=0.31; *p*<0.01). The correlation was weak between the motor domain of the ESDI and the fine motor domain of the BSID (*r*=0.22; *p*<0.05). A table with the results by age group is presented in Supplementary File I.

**Figure 2:**
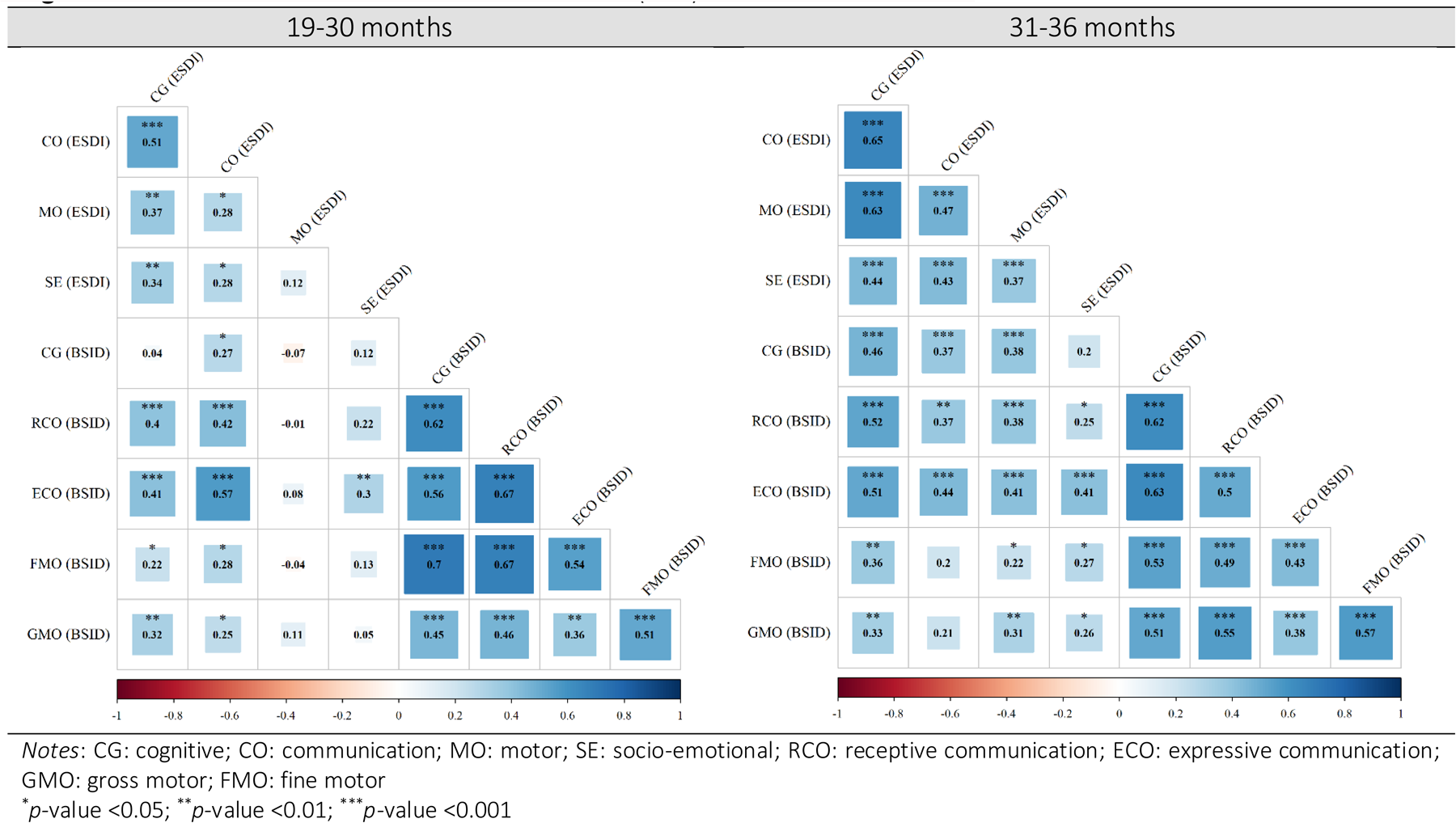
correlation matrix between ESDI and BSID (raw) domains

### 3.4. Relationship between ESDI and BSID scores with maternal education and household wealth

The ESDI domains cognitive, communication, and motor skills as well as the BSID domains cognitive, receptive communication, expressive language, fine motor, and gross motor skills were significantly and positively associated with maternal education. No significant correlation was found between the socio-emotional domain of the ESDI and maternal education. In the household wealth classification, only significant associations were found with the ESDI domain motor skills and the BSID domains cognitive an expressive language skills (Table 4).

**Table 4.** Relationship between ESDI and BSID (raw scores) with maternal education and household wealth (all ages combined)

| Maternal education<sup>a</sup> | Wealth classification<sup>a,b</sup> |
| --- | --- |
| Coefficient (95% IC) | Coefficient (95% IC) |

| ESDI scores |  |  |
| --- | --- | --- |
| Cognitive | 1.55 (0.61, 2.49)** | 32.8 (-6.85, 72.5) |
| Communication | 1.41 (0.26, 2.56)* | 55.49 (-2.85, 113.83) |
| Motor | 0.93 (0.18, 1.68)* | 26.50 (1.44, 51.57)* |
| Socio-emotional | 0.22 (-0.27, 0.71) | 7.10 (-13.96, 28.16) |
| BSID (raw) scores |  |  |
| Cognitive | 0.38 (0.21, 0.55)*** | 8.38 (3.07, 13.69)** |
| Receptive language | 0.25 (0.05, 0.46)* | 5.11 (-1.11, 11.33) |
| Expressive language | 0.30 (0.10, 0.51)** | 8.88 (2.69, 15.06)** |
| Fine motor | 0.18 (0.04, 0.39)* | 4.50 (-2.49, 1.50) |
| Gross motor | 0.34 (0.12, 0.55)** | 2.42 (-5.83, 10.67) |

<sup>a</sup>GEE regressions adjusted for child's sex, child's age and within cluster correlation
<sup>b</sup>According to the Young Lives Wealth Index (Briones, 2017).
\**p*-value <0.05; \*\**p*-value <0.01; \*\*\**p*-value <0.001

### 4. Discussion

In this study, we investigated the internal consistency and concurrent validity of the Peruvian Infant Development Scale (ESDI) screening tool used by the Peruvian National early child development programme (PNCM) for children under 36 months of age. The PNCM developed the ESDI to assess the developmental status of children in the complex cultural contexts of Peru. We compared ESDI scores with those obtained using the internationally validated Bayley Scale of Infant Toddler Development (BSID) tool, which is considered the gold standard for measuring children’s developmental status, in two age ranges: 19-30 and 31-36 months. To our knowledge, this is one of the first studies to compare the ESDI tool with an internationally validated screening tool. The study examined 167 children who had previously participated in a trial conducted in high-altitude rural Andean Peru.

The ESDI instrument (all ages combined) showed good internal consistency and reliability (α=0.75). The item-test correlations of the ESDI to assess the internal structure of the instruments were within the good ranges indicated, although some differences were observed between the developmental domains. The item-test correlations of the socio-emotional and motor domains of the ESDI were lower than those of the cognitive and communication domains. This suggests that it may be difficult to discriminate between high and low performers in these domains. The correlation matrix confirmed these results, showing weak correlations between the motor domain of the ESDI and the fine and gross motor domains of the BSID at 19-30 and 31-36 months. The BSID item-test correlations showed no noticeable differences between the developmental domains.

The ESDI was found to have moderate concurrent validity in the communication domain at 19-30 months and in the communication and cognitive domains at 31-36 months. Correlations between the motor domains of the ESDI and BSID were weak in both age ranges. For this reason, particular attention should be paid to the assessment of motor skill with the ESDI, especially in children aged 19-30 months. The concurrent validity of the ESDI is comparable to that of other similar instruments. Three multidimensional screening instruments validated in Colombia showed age-dependent improvements in concurrent validity for children over 30 months of age in all developmental domains except gross motor skills (Rubio-Codina et al., 2016).

Some factors may contribute to the unequal psychometric values observed for the ESDI instrument at 19-30 months of age. The ESDI was administered by fieldworkers rather than professionals, and the assessment conditions were not standardised (e.g., some assessments were conducted in households, while others took place in locations where there were more distractions, such as courtyards). In addition, as the children had participated in a trial for a year, it was difficult to divide the group of 19-30 months into smaller subgroups. This would have been interesting to better explain how the concurrent validity of ESDI domains changed over time.

In addition, we found that few BSID domains were associated with household wealth compared to other studies conducted in China and Colombia (Rubio-Codina et al., 2016; Yue et al., 2019). We believe that this difference can be explained by the homogeneity of our sample, as only poor and vulnerable families were included in the trial.

Our study has some limitations. Due to a lack of time constraints, we only carried out one assessment per instrument. A test-retest evaluation to assess reliability over time was not possible. In addition, exploratory and confirmatory factor analyses would further corroborate our results. Unfortunately, we were unable to enter responses for each item (domains only) due to logistical limitations.

Finally, it is possible that the psychometric properties of the ESDI were influenced by the fact that local staff and not PNCM staff conducted the assessments. However, our study shows that the ESDI had good psychometric properties, even when administered by non-experts. This makes the ESDI a valuable alternative to the BSID and supports the integration of the ESDI tool into the PNCM expansion in Peru.

Overall, the ESDI can be used to measure the developmental status of children in rural high-altitude Andean Peru, especially children over 30 months of age. As expected, the BSID had better psychometric properties. However, the ESDI is free, brief, and can be easily administered without qualified personnel or expensive materials. The ESDI tool therefore offers a practical and cost-effective alternative for remote areas in Peru with limited human and logistical resources.

## 5. Conclusion

We analysed the internal consistency and concurrent validity of the Peruvian Infant Development Scale (ESDI) screening tool to measure the developmental status of children in high-altitude rural Andean Peru. The Bayley Scales of Infant Toddler Development (BSID) instrument was used to compare the results. Despite the higher accuracy of the BSID, the ESDI showed adequate internal consistency, reliability, and concurrent validity for most developmental domains, especially for children over 30 months of age. In addition, the ESDI and BSID scores were equally correlated with maternal education and household wealth. According to these results, the ESDI is a suitable alternative tool for assessing the developmental status of children in rural areas where human, logistical, and economic resources are limited.

## Data Availability

The data and materials underlying this article will be shared on reasonable request to the corresponding author.

