## Supplemental Table 1 for "Internal consistency and concurrent validity of the Peruvian Infant Development Scale tool in high-altitude rural Andean Peru"

**Table S1**. Correlation matrix between ESDI and BSID (raw) scores by age group

| | | BSID domains | | | | |
|---|---|---|---|---|---|---|
| | | Cognitive | Receptive language | Expressive language | Fine motor | Gross motor |
| | ESDI domains | | | | | |
| 19-30 months[a] | Cognitive | **0.04 (-0.14, 0.22)** | 0.40 (0.21, 0.59)$^{***}$ | 0.41 (0.24, 0.59)$^{***}$ | 0.22 (0.02, 0.42)$^{*}$ | 0.32 (0.16, 0.48) $^{***}$ |
| | Communication | 0.27 (0.02, 0.51)$^{**}$ | **0.42 (0.26, 0.58)**$^{***}$ | **0.57 (0.38, 0.75)**$^{***}$ | 0.28 (0.06, 0.49)$^{*}$ | 0.25 (0.06, 0.44)$^{**}$ |
| | Motor | -0.07 (-0.24, 0.10) | -0.00 (-0.23, 0.21) | 0.08 (-0.16, 0.32) | **-0.04 (-0.22, 0.14)** | **0.11 (-0.06, 0.28)** |
| | Socio-emotional | 0.12 (-0.06, 0.31) | 0.22 (0.03, 0.40)$^{*}$ | 0.30 (0.09, 0.52)$^{**}$ | 0.13 (-0.02, 0.28) | 0.05 (-0.16, 0.26) |
| | | BSID domains | | | | |
| | | Cognitive | Receptive language | Expressive language | Fine motor | Gross motor |
| | ESDI domains | | | | | |
| 31-36 months[a] | Cognitive | **0.46 (0.32, 0.60)**$^{***}$ | 0.52 (0.40, 0.65)$^{***}$ | 0.51 (0.39, 0.62)$^{***}$ | 0.36 (0.18, 0.53)$^{***}$ | 0.33 (0.17, 0.49)$^{***}$ |
| | Communication | 0.36 (0.18, 0.53)$^{***}$ | **0.36 (0.18, 0.53)**$^{**}$ | **0.44 (0.30, 0.58)**$^{***}$ | 0.20 (0.06, 0.35)$^{**}$ | 0.21 (0.05, 0.37)$^{*}$ |
| | Motor | 0.38 (0.22, 0.53)$^{***}$ | 0.38 (0.23, 0.54)$^{***}$ | 0.41 (0.28, 0.55)$^{***}$ | **0.22 (0.03, 0.40)**$^{*}$ | **0.31 (0.13, 0.49)**$^{**}$ |
| | Socio-emotional | 0.20 (0.00, 0.40) | 0.25 (0.04, 0.46)$^{*}$ | 0.41 (0.26, 0.57)$^{***}$ | 0.27 (0.07, 0.46)$^{**}$ | 0.26 (0.09, 0.44)$^{**}$ |
| | | BSID domains | | | | |
| | ESDI domains | Cognitive | Receptive language | Expressive language | Fine motor | Gross motor |
| All ages combined[a] | Cognitive | **0.37 (0.25, 0.50)**$^{***}$ | 0.53 (0.42, 0.65)$^{***}$ | 0.53 (0.43, 0.64)$^{***}$ | 0.43 (0.31, 0.54)$^{***}$ | 0.43 (0.30, 0.55)$^{***}$ |
| | Communication | 0.44 (0.29, 0.59)$^{***}$ | **0.51 (0.39, 0.64)**$^{***}$ | **0.60 (0.49, 0.70)**$^{***}$ | 0.40 (0.27, 0.52)$^{***}$ | 0.37 (0.24, 0.50)$^{***}$ |
| | Motor | 0.09 (-0.04, 0.22) | 0.12 (-0.02, 0.27) | 0.19 (0.05, 0.34)$^{**}$ | **0.07 (-0.07, 0.20)** | **0.16 (0.02, 0.30)** $^{*}$ |
| | Socio-emotional | 0.30 (0.16, 0.44)$^{***}$ | 0.37 (0.23, 0.50)$^{***}$ | 0.45 (0.31, 0.59)$^{***}$ | 0.34 (0.23, 0.46)$^{***}$ | 0.29 (0.14, 0.43)$^{***}$ |

[a]Pearson correlations (with IC 95%) and bootstrapping methods of 1,000 replications.
$^{*}$*p*-value <0.05; $^{**}$*p*-value <0.01; $^{***}$*p*-value <0.001
